# Incorporating Algorithmic Uncertainty into a Clinical Machine Deep Learning Algorithm for Urgent Head CTs

**DOI:** 10.1101/2022.07.19.22277808

**Authors:** Byung C. Yoon, Stuart R. Pomerantz, Nathaniel D. Mercaldo, Swati Goyal, Eric L’Italien, Michael H. Lev, Karen Buch, Bradley R. Buchbinder, John W. Chen, John Conklin, Rajiv Gupta, George J. Hunter, Shahmir M. Kamalian, Hillary R. Kelly, Otto Rapalino, Sandra P. Rincon, Javier M. Romero, Julian He, Pamela W. Schaefer, Synho Do, R. Gilberto González

## Abstract

Machine learning (ML) algorithms to detect critical findings on head CTs may expedite patient management. Most ML algorithms for diagnostic imaging analysis utilize dichotomous classifications to determine whether a specific abnormality is present. However, imaging findings may be indeterminate, and algorithmic inferences may have substantial uncertainty. We incorporated awareness of uncertainty into an ML algorithm that detects intracranial hemorrhage or other urgent intracranial abnormalities and evaluated prospectively identified, 1000 consecutive noncontrast head CTs assigned to Emergency Department Neuroradiology for interpretation. The algorithm classified the scans into high (IC+) and low (IC-) probabilities for intracranial hemorrhage or other urgent abnormalities. All other cases were designated as No Prediction (NP) by the algorithm. The positive predictive value for IC+ cases (N = 103) was 0.91 (CI: 0.84-0.96), and the negative predictive value for IC-cases (N = 729) was 0.94 (0.91-0.96). Admission, neurosurgical intervention, and 30-day mortality rates for IC+ was 75% (63-84), 35% (24-47), and 10% (4-20), compared to 43% (40-47), 4% (3-6), and 3% (2-5) for IC-. There were 168 NP cases, of which 32% had intracranial hemorrhage or other urgent abnormalities, 31% had artifacts and postoperative changes, and 29% had no abnormalities. An ML algorithm incorporating uncertainty classified most head CTs into clinically relevant groups with high predictive values and may help accelerate the management of patients with intracranial hemorrhage or other urgent intracranial abnormalities.

## Background

A patient with an urgent neurological abnormality requires prompt imaging. If the etiology is hemorrhage, immediate evacuation or treatment of an aneurysm may be lifesaving. If the cause is an ischemic stroke from a large vessel occlusion, thrombectomy can result in a favorable outcome. There are many additional causes of urgent neurological deficits including masses, infections, inflammatory diseases, and acute hydrocephalus for which prompt intervention could markedly decrease morbidity and mortality. Rapid identification of such a finding is critical for triaging patients to appropriate management. CT is commonly the first imaging method employed in this situation, and computer-based image evaluation tools that rapidly classify and triage CT scans would be valuable.

The application of artificial intelligence (AI) in neuroimaging has expanded with the potential to accelerate the accurate diagnosis of intracranial hemorrhage or other urgent intracranial abnormalities. Many machine-learning (ML) algorithms rely on dichotomous classification schemes where an algorithm determines the presence or absence of specific intracranial abnormalities such as intracranial hemorrhage, large vessel occlusion, or metastasis.(1-4) However, abnormal findings on neuroimaging are not always definitive. For instance, a streak artifact on CT may mimic intracranial hemorrhage. Uncertainties can also be introduced at different stages in the development of ML algorithms for various reasons including suboptimal input data, weak supervision, as well as inter- and intra-observer variability.(5) With the expansion of real-life applications of various ML algorithms and the growing need for safety, the recognition and appropriate handling of uncertainty is becoming critical.(6)(7)

Clinical ambiguity is also common. For instance, stroke symptoms including a facial droop and speech difficulty may not only result from early ischemia but also from acute intracranial hemorrhage or a mass lesion. An appropriate neurological assessment may not be feasible in patients who are obtunded from various causes such as ischemia, infection, and inflammation. The clinical issue at hand is therefore complex, and more than one ML algorithm may be needed for adequate characterization of the head CT, including an algorithm that rapidly identifies a major abnormality such as hemorrhage, mass lesions, and other stroke mimics as well as another algorithm that quickly assesses the likelihood of ischemic injury.

In this study, our aim was to evaluate an ML tool that classifies non-contrast CT scans into three categories based on the probability of the presence of an intracranial hemorrhage or other urgent intracranial abnormality other than acute ischemia. We assessed the impact of this classification in predicting clinical outcomes including hospital admission, neurosurgical intervention, and 30-day mortality.

## Materials and Methods

### Deep Learning Algorithm

The algorithm employed was derived from an intracranial hemorrhage detection and subtype classification system first created by finetuning four ImageNet-pretrained - deep convolutional neural networks (DCNNs)—VGG16,(8) ResNet-50,(9) Inception-v3, (10) and Inception-ResNet-v2, (11) with ICH training data obtained from the imaging archives of our institution.(12) Specifically, 5 mm thick two-dimensional (2D) axial images from 904 non-contrast head CTs were labeled by 5 subspecialty-trained US board-certified neuroradiologists as to the absence or presence of intracranial hemorrhage as well as all identifiable subtypes (intraparenchymal, intraventricular, subdural, epidural, subarachnoid). These studies were randomly divided into training and validation sets and additional retrospectively and prospectively collected data sets were used for testing. Finetuning of the DCNNs with this labeled data occurred after the last fully-connected layers were replaced with three consecutive layers containing a global average pooling (GAP)(13) layer, a fully-connected layer, and an element-wise sigmoid layer. All models were optimized using a mini-batch stochastic gradient descent with Nesterov momentum(14) with a batch size of 64 to maximize GPU utilization. We used a weight decay of 5×10^−5^ and a base learning rate of 0.001, decayed by 0.1 three times when the validation loss plateaus. Computations were conducted on an NVIDIA DevBox equipped with four TITAN X GPUs with 12GB of memory per GPU, and all deep learning models were implemented using Keras (version 2.1.2, http://keras.io) with a Tensorflow(15) backend (version 1.3.0). The model provided a probability on a slice-by-slice basis of the presence of intraparenchymal hemorrhage, interventricular hemorrhage, subdural hematoma, epidural hematoma, and subarachnoid hemorrhage. This multi-label classification task was reformulated into a binary classification of intracranial hemorrhage as positive if one or more of the subtype outputs are positive and negative if not. The binary cross entropy loss function was weighted by the ratios of positive and negative instances for each class label, in a similar fashion as described previously(16, 17). An ensemble of the four models was created using unweighted averaging such that the final probability is defined as an average of probabilities predicted by the four models. We also applied the Z directional moving average window method to determine the results of the target image by considering the results of the images above and below the Z-axis. This method is more accurate than the 2D method and has better memory usage and algorithm speed than the 3D method. The model assigned for each slice of a CT scan, a probability of 0 to 1 for the presence of intracranial hemorrhage. Complete details are described by Lee et al.(12)

### Adapting the Algorithm for Clinical Use

The algorithm underwent a preliminary evaluation using ED neuroradiology head CTs acquired over a period of one month. Review of the results revealed that the algorithm identified as abnormal significant intracranial abnormalities other than ICH. We deduced that this was a form of transfer learning and a valuable feature of the algorithm. We also observed that when a study had 3 or more slices with probabilities of at least 0.9, ICH or other significant intracranial abnormality was very likely. We also noted that if a study contained just 1 or no images with probability greater or equal to 0.6, ICH or other abnormality was very unlikely. For our prospective study we classified studies with 3 or more slices with probabilities of 0.9 or greater as IC+; studies with all slices except one having probabilities of 0.6 or less were classified as IC-. All other studies were given the designation of NP indicating no prediction by the algorithm.

### Imaging Test Data Collection

This study was approved by the institutional review board (IRB). Prospectively, we identified 1000 consecutive non-contrast head CTs assigned for review by the emergency department neuroradiology service. Most of the studies were non-contrast head CT, but some were performed in conjunction with CT angiography. Regardless of the type of the study, only the axial, 5-mm, noncontrast, standard reconstruction kernel, head CT images were evaluated. Most of the studies (N = 953) were acquired in the emergency department or inpatient CT scanners (GE Discovery CT750 HD, GE LightSpeed VCT, Siemens SOMATOM Definition Edge). The remaining 47 studies were performed on the outpatient scanners (Philips IQon -Spectral CT, GE Discovery CT750 HD).

The clinical information was derived from electronic medical records. In addition to patients’ demographic data, the rates of admission, neurosurgical intervention, and 30-day mortality were also collected. Neurosurgical interventions included craniotomy, craniectomy, cranioplasty, ventricular catheter placement or removal, biopsy, and interventional neuroradiology procedures. If a patient had multiple imaging exams during the same admission, it was counted as a single admission. There were 4 patients who were discharged and re-admitted subsequently during the data collection. Their admissions were counted as two separate admissions.

### Imaging Data Analysis and Clinical Information

Each CT scan classified by the algorithm was compared to the formal clinical interpretation by neuroradiologists with certificates of added qualification (CAQ), which served as the ground truth. The clinical interpretations were made without the knowledge of the ML algorithm results. The presence of intracranial hemorrhage or other urgent intracranial abnormalities was recorded and compared with the algorithm results.

### Statistical Analysis

Descriptive summaries were computed for the overall cohort. Continuous variables were summarized as the median and interquartile range (IQR: 25^th^ and 75^th^ quantile) and categorical variables as percentages.

Separate generalized estimating equations, with an independence correlation structure (GEE-I), were used to quantify the association between classification (IC+, IC-, NP) and each imaging finding (acute intracranial hemorrhage, mass lesion, artifact, post-op changes, acute/subacute infarcts, encephalomalacia, miscellaneous, and normal imaging) and each outcome (hospital admission, neurosurgical intervention, and 30-day mortality). Linear combinations of parameter estimates were computed to summarize the prevalence of each imaging finding and the occurrence of each outcome by classification group as well as all pairwise comparisons (outcomes only) between classification groups along with their associated confidence intervals and P values. Similarly, GEE-I estimates were constructed to estimate predictive values while acknowledging the possibility of multiple admissions and scans per subject. All analyses were performed using R 4.1.1 (R: the R project for statistical computing; www.r-project.org) and the geepack library.(18)

## Results

One thousand consecutive head CT scans were performed on 857 patients. Some patients had multiple studies during the data collection period. Of these 857 subjects, 761 had a single scan, while 66 had two scans, and 33 had three or more scans. There were 423 female (49.4%) and 434 male (50.6%) patients. The median age was 65 years (IQR: 50-77). The overall hospital admission rate was 47.1% (CI: 43.8-50.4) which is higher than the ∼30% admission rate of all patients evaluated in our institution’s emergency department. Neurosurgical interventions, including craniotomies and endovascular procedures, were performed in 7.7% (6.1-9.7) and the 30-day mortality was 4.4% (3.2-6.0).

Of the 1000 scans, 10.3% were classified by the algorithm as IC+ (high probability of an intracranial hemorrhage or other urgent intracranial abnormality), and 72.9% were classified as IC- (low probability of an ICH or other urgent intracranial abnormality). Studies classified as No Prediction (NP) constituted 16.8% of the 1000 scans. Table 1 lists the imaging findings tabulated for IC+ and IC- cases.

**Table 1.**
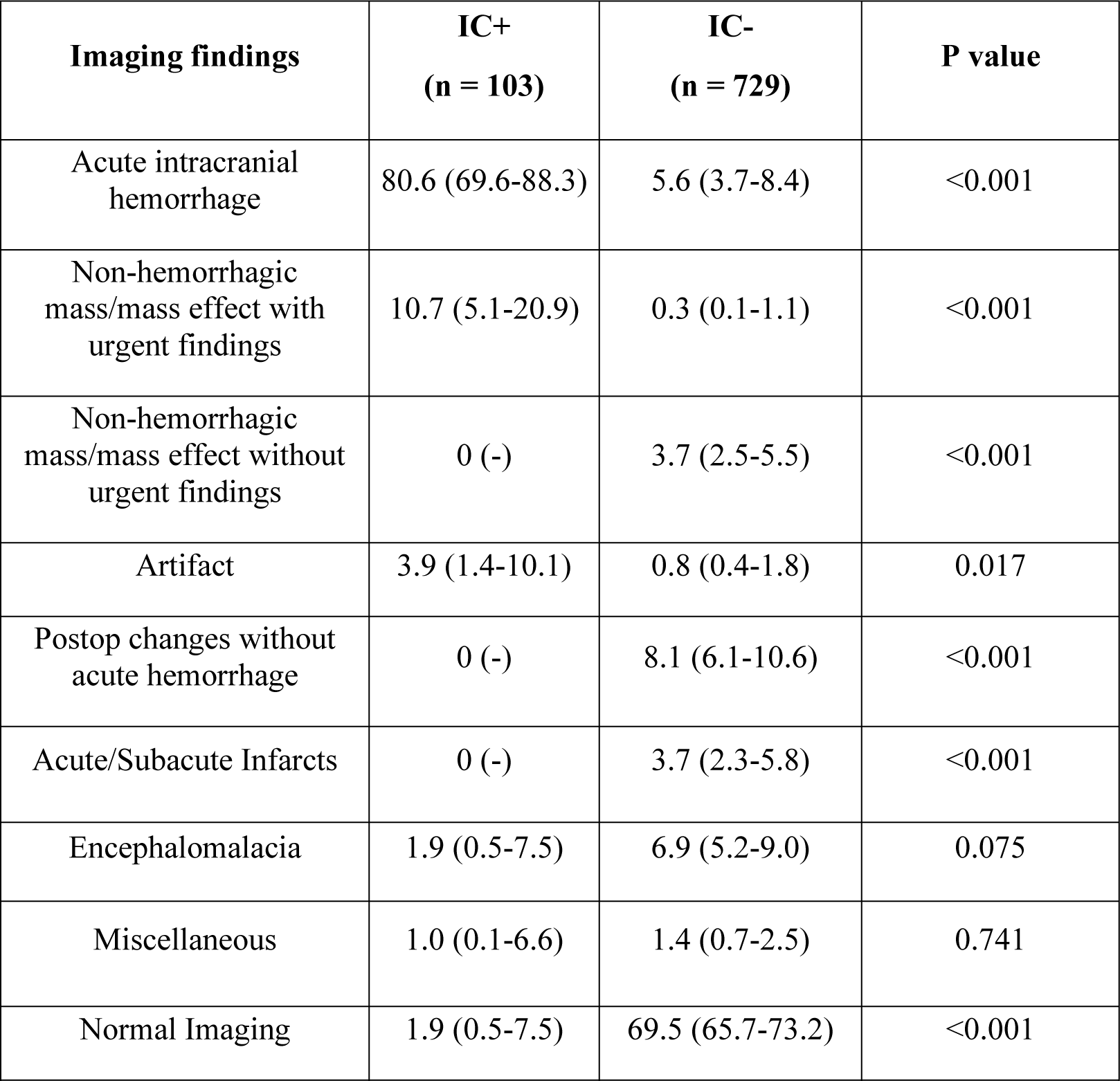
Neuroimaging Abnormality Distribution by Classification. The proportion in percent (95% confidence interval) of major intracranial abnormalities for studies classified as IC+ and IC-. Intracranial hemorrhage includes acute hemorrhage associated with aneurysmal rupture, posttraumatic, hemorrhagic infarcts, hemorrhagic mass, and/or postoperative changes. Urgent findings associated with mass/mass effect includes herniation, ventricular effacement, and/or ventricular entrapment from various etiologies including neoplasm, infection, inflammation, and cerebral edema. Postoperative changes include findings such as resection cavity or pneumocephalus from craniotomy without acute intracranial hemorrhage or substantial mass effect. Acute/subacute infarcts include non-hemorrhagic ischemic infarction. Miscellaneous includes ventriculomegaly, chronic subdural collections, and hygromas. All estimates (main effects: percentage, 95% CI, comparisons: odds ratios, 95% CI, P values) are based on separate generalized estimating equations using an independent correlation structure.

| **Imaging findings** | **IC+**<br>**(n = 103)** | **IC-**<br>**(n = 729)** | **P value** |
| --- | --- | --- | --- |
| Acute intracranial hemorrhage | 80.6 (69.6-88.3) | 5.6 (3.7-8.4) | <0.001 |
| Non-hemorrhagic mass/mass effect with urgent findings | 10.7 (5.1-20.9) | 0.3 (0.1-1.1) | <0.001 |
| Non-hemorrhagic mass/mass effect without urgent findings | 0 (-) | 3.7 (2.5-5.5) | <0.001 |
| Artifact | 3.9 (1.4-10.1) | 0.8 (0.4-1.8) | 0.017 |
| Postop changes without acute hemorrhage | 0 (-) | 8.1 (6.1-10.6) | <0.001 |
| Acute/Subacute Infarcts | 0 (-) | 3.7 (2.3-5.8) | <0.001 |
| Encephalomalacia | 1.9 (0.5-7.5) | 6.9 (5.2-9.0) | 0.075 |
| Miscellaneous | 1.0 (0.1-6.6) | 1.4 (0.7-2.5) | 0.741 |
| Normal Imaging | 1.9 (0.5-7.5) | 69.5 (65.7-73.2) | <0.001 |

The algorithm classified 103 cases as IC+. Examples of IC+ scans are shown in Figure 1. Over 80% of these patients had intracranial hemorrhages, either primary or secondary to underlying lesions identified on the non-contrast CT or associated examinations.

**Figure 1.**
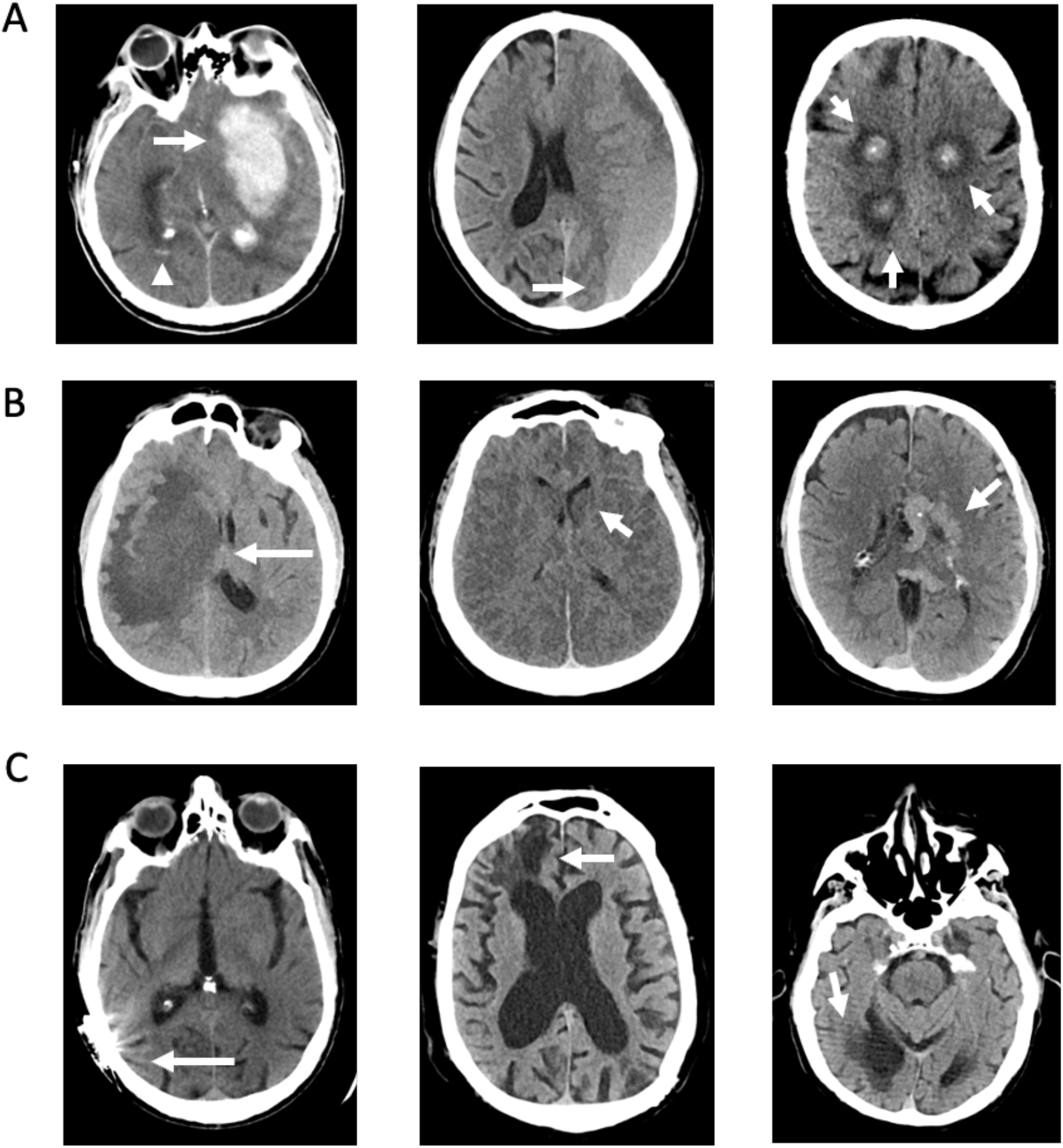
IC+ Cases. **A:** Examples of IC+ cases with acute intracranial hemorrhages. The left panel shows a large intraparenchymal hematoma centered in the left basal ganglia (arrow) as well as intraventricular hemorrhages within the bilateral occipital horns (arrowhead). The middle panel shows a large left holohemispheric subdural hematoma with rightward subfalcine herniation. The right panel shows multifocal, hemorrhagic lung cancer metastases. **B:** Examples of nonhemorrhagic IC+ cases. The left panel shows a large area of confluent hypoattenuation involving the right hemisphere, which was subsequently found to be toxoplasmosis in a patient with human immunodeficiency virus (HIV) and acquired immunodeficiency syndrome (AIDS). The middle panel shows diffuse reversal of gray-white differentiation as well as partial effacement of the ventricles, most consistent with diffuse cerebral edema from a severe hypoxic-ischemic injury. For instance, the left caudate head has an abnormally hypodense appearance compared to the adjacent internal capsule. The right panel shows a large, left hemispheric, Spetzler Martin grade 5 arteriovenous malformation. **C:** Examples of false positive IC+ cases. The left panel shows streak artifact from metallic hardware. The middle panel shows encephalomalacia of the right frontal lobe. The right panel shows an old right posterior cerebral artery territory infarction.

Non-hemorrhagic masses or lesions causing substantial mass effect were the second most common findings in over 10% of cases. Several cases with urgent pathology including diffuse cerebral edema and large vascular malformation were also classified as IC+. There were 9 IC+ false positives. There was only a single normal study that was classified as IC+ with the erroneous classification thought to be secondary to residual intracranial vascular enhancement from an earlier exam performed with intravenous contrast.

Among the IC+ cases, there were 94 true positive and 9 false positive cases for major intracranial abnormality, resulting in a positive predictive value 0.91 (0.84-0.96). Patients with IC+ scans had a very high rate of admission, 74.6% (62.5-83.8). The rates of neurosurgical intervention, 34.9% (24.2-47.4), and 30-day mortality, 9.5% (4.3-19.6), were also high. These outcomes were significantly greater in IC+ patients compared to IC- patients (Table 2).

**Table 2.** Clinical Outcomes by Classification. All estimates (main effects: percentage, 95% CI, comparisons: odds ratios, 95% CI, P values) are based on separate generalized estimating equations using an independent correlation structure.

| Outcome | Overall | Classification |  | Comparison |
| --- | --- | --- | --- | --- |
|  |  | IC+<br>Percent<br>(95% CI) | IC-<br>Percent<br>(95% CI) | IC+ vs IC-<br>Odds Ratio<br>(95% CI), P value |
| Admission | 47.1<br>(43.8-50.4) | 74.6<br>(62.5-83.8) | 43.3<br>(39.6-47.1) | 3.85<br>(2.14-6.91), P <0.001 |
| Neurosurgical Intervention | 7.7<br>(6.1-9.7) | 34.9<br>(24.2-47.4) | 4.3<br>(3.0-6.1) | 11.90<br>(6.29-22.50), P <0.001 |
| 30-Day Mortality | 4.4<br>(3.2-6.0) | 9.5<br>(4.3-19.6) | 3.0<br>(1.9-4.6) | 3.42<br>(1.32-8.86), P = 0.011 |

The findings on the 9 false positive IC+ scans are listed in Table 3. The false positive cases included 4 with artifacts, 4 with large areas of encephalomalacia and one from a patient that received intravenous contrast during an earlier exam.

**Table 3.**
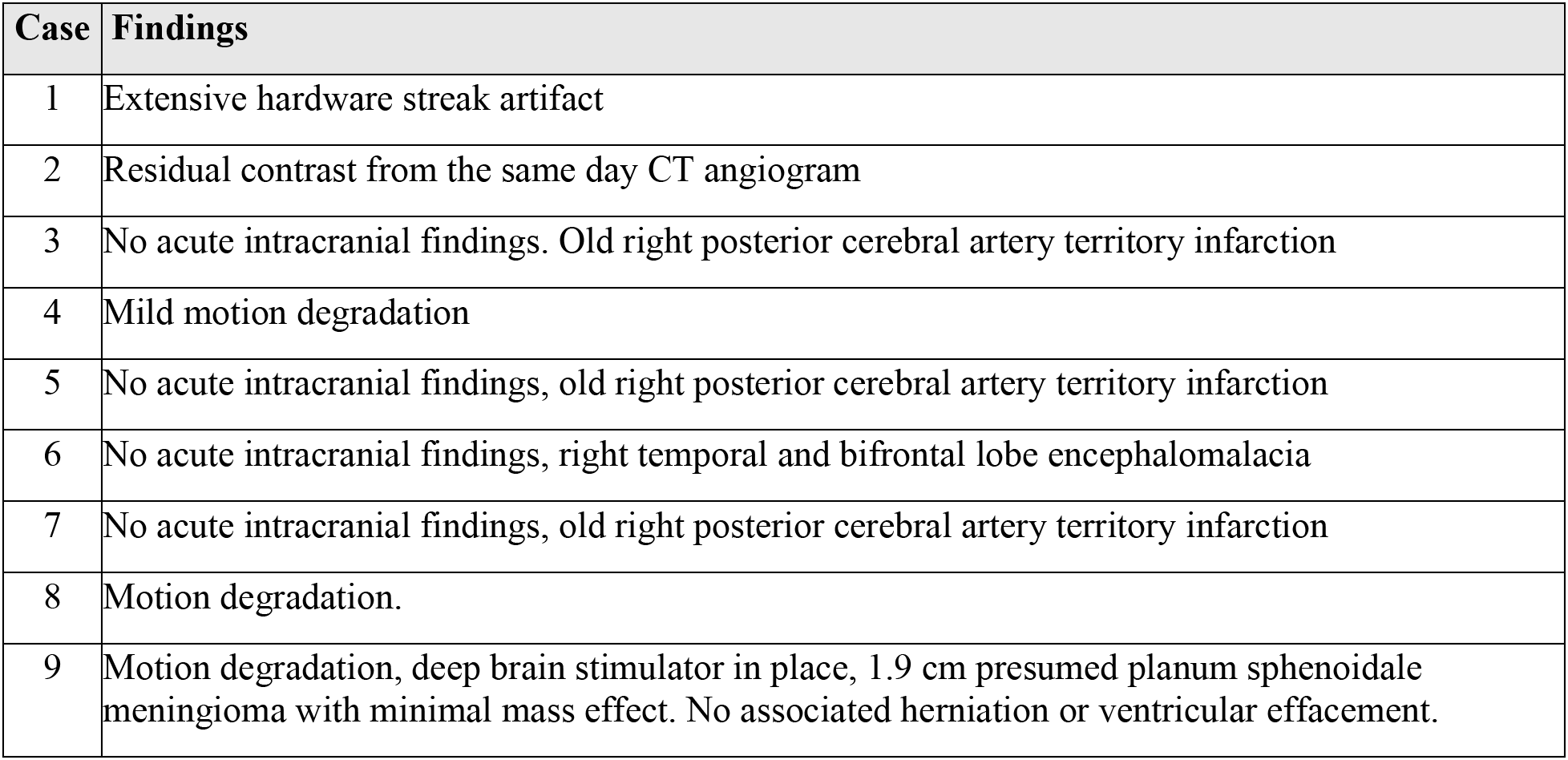
False positive IC+ cases.

| Case | Findings |
| --- | --- |
| 1 | Extensive hardware streak artifact |
| 2 | Residual contrast from the same day CT angiogram |
| 3 | No acute intracranial findings. Old right posterior cerebral artery territory infarction |
| 4 | Mild motion degradation |
| 5 | No acute intracranial findings, old right posterior cerebral artery territory infarction |
| 6 | No acute intracranial findings, right temporal and bifrontal lobe encephalomalacia |
| 7 | No acute intracranial findings, old right posterior cerebral artery territory infarction |
| 8 | Motion degradation. |
| 9 | Motion degradation, deep brain stimulator in place, 1.9 cm presumed planum sphenoidale meningioma with minimal mass effect. No associated herniation or ventricular effacement. |

Most patients (N = 729) had scans classified as IC-, and examples are shown in Figure 2. The negative predictive value was 0.94 (0.91-0.96).

**Figure 2.**
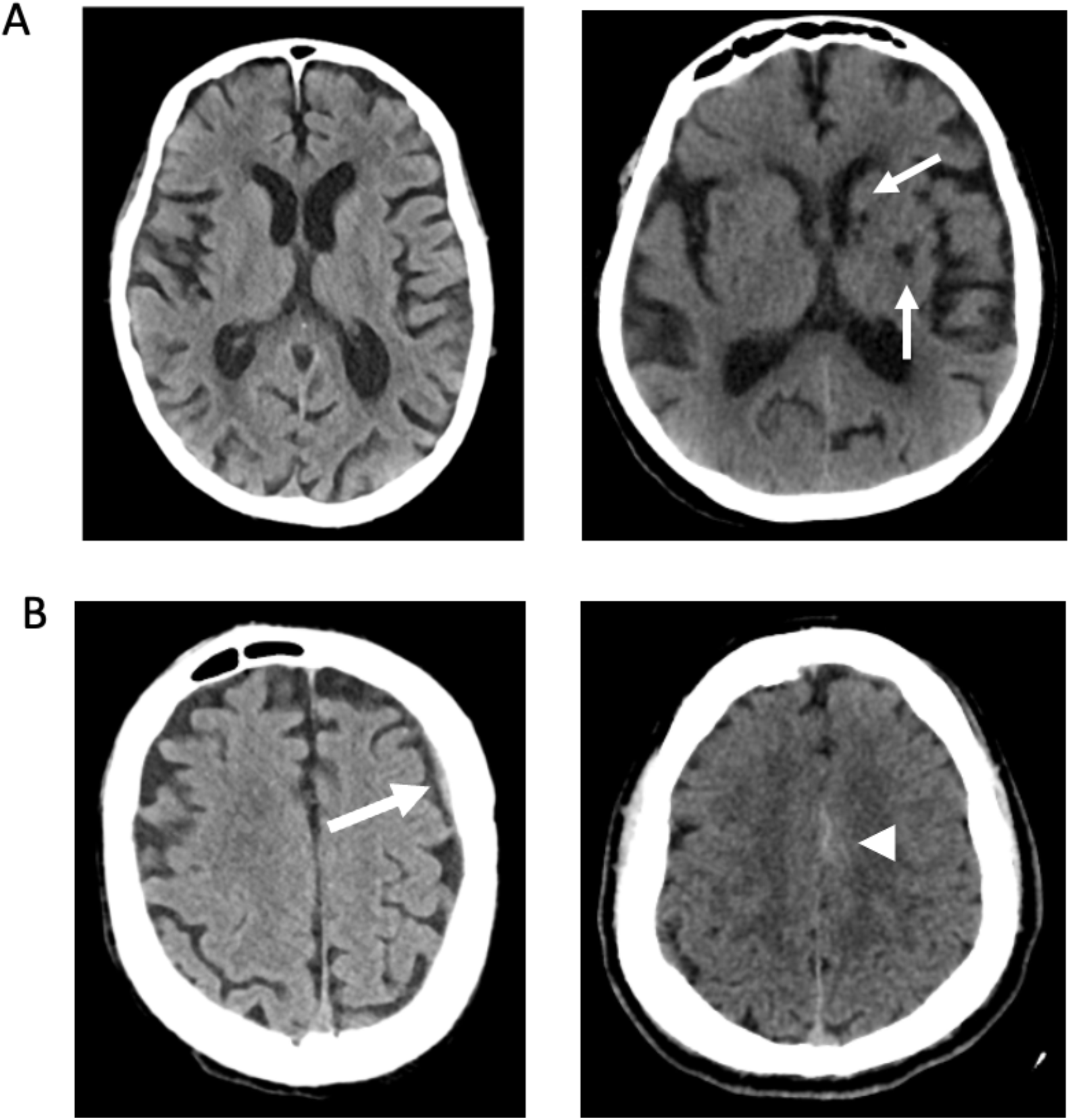
IC- cases. **A:** Examples of true negative IC- cases. The left panel shows a normal noncontrast head CT. The right panel shows small old infarcts in the left basal ganglia. **B:** Examples of false negative IC- cases. The left panel shows a small isodense subdural hematoma along the left frontal convexity. The right panel shows trace subarachnoid hemorrhage along the medial aspect of the left frontal lobe.

The rates of admission, neurosurgical intervention, and 30-day mortality for the IC- group were substantially lower than those in the IC+ group (Table 1). There were 686 true negative cases, and they had no abnormal findings or small chronic abnormalities (Figure 2). There were 43 scans that were classified as IC- that were false negatives (Table 4). Nearly all, 39 of 41, of the false negative scans had only very small areas of hemorrhage, petechial intraparenchymal or trace extra-axial in nature, without appreciable mass effect. (Figure 2). The remaining 2 false-negative cases had mass lesions in the cerebellum with partial effacement of the fourth ventricles.

**Table 4.** False negative IC- cases.

| Case | Findings |
| --- | --- |
| 1 | Small bilateral SDH |
| 2 | Small bilateral SDH |
| 3 | Right SDH measuring up to 6 mm in thickness |
| 4 | Small volume acute SAH over the left frontal convexity and possibly in the dependent portion of the left sylvian fissure. |
| 5 | Trace SAH along the left frontal lobe |
| 6 | Left hypodense to isodense SDH |
| 7 | Trace scattered SAH |
| 8 | Trace SDH |
| 9 | Right thalamic possible ICH |
| 10 | Right thalamic IPH |
| 11 | Small volume SAH along the inferior left temporal lobe extending to the left temporo-occipital junction and trace SDH along the left tentorial leaflet |
| 12 | Thin linear hyperattenuation along the right tentorial leaflet, consistent with a small SDH |
| 13 | Small left frontal and parietal SDH |
| 14 | Small scattered foci of extra-axial hemorrhage along the bilateral frontal convexities and falx |
| 15 | Mildly hyperdense 2 mm extra-axial collection concerning for small SDH |
| 16 | Small foci of extra-axial hemorrhage along the frontal convexities. |
| 17 | Small right frontal extra-axial hematoma |
| 18 | Bifrontal extra-axial hyperdense foci, SDH vs myeloid sarcoma in the setting of AML |
| 19 | Small right frontal extra-axial hematoma. |
| 20 | Multicompartmental ICH (isodense right SDH, evolving predominantly hypodense R IPH) |
| 21 | Small hemorrhagic contusions at the left middle frontal gyrus with surrounding vasogenic edema, with a possible component of subarachnoid hemorrhage. |
| 22 | 2 mm left parietal convexity SDH |
| 23 | Small left parafalcine SAH |
| 24 | Left frontal convexity and right parafalcine SDH |
| 25 | Left frontal contusion and trace extra-axial hemorrhage |
| 26 | Possible subacute infarct with hemorrhagic transformation in the posterior left temporal lobe |
| 27 | Minimal tract hemorrhage along the previously seen course of ventricular catheter embedded within the corpus callosum splenium. |
| 28 | Small hemorrhagic contusion |
| 29 | Less dense appearance of extra-axial foci along the bilateral frontal convexities, likely reflecting evolving blood products |
| 30 | Evolving posterior left temporal subacute infarct with a small degree of hemorrhagic transformation |
| 31 | Bilateral SDH |
| 32 | Trace right parietal SAH |
| 33 | Small focus of acute subdural hematoma along the right aspect of the posterior falx cerebri |
| 34 | Subdural collection with acute blood products or granulation tissue |
| 35 | SAH |
| 36 | Mass effect in the left cerebellar hemisphere with partial effacement of the fourth ventricle |
| 37 | Left cerebellar metastasis with partial effacement of the fourth ventricle |
| 38 | SAH |
| 39 | SAH |
| 40 | Stable small left parietal hematoma, possible trace left parietal SAH |
| 41 | Trace SAH in the suprasellar and interpeduncular cisterns and small IVH |
| 42 | Small volume traumatic SAH |
| 43 | Small volume traumatic SAH |

SDH = subdural hematoma, SAH = subarachnoid hemorrhage, ICH = intracranial hemorrhage, IVH = intraventricular hemorrhage, IPH = intraparenchymal hematoma, AML = acute myeloid leukemia

The NP patient group included 168 scans (Figure 3). Many of the patients had urgent abnormalities including 41 with intracranial hemorrhage, and 13 with mass lesions or substantial mass effect.

**Figure 3.**
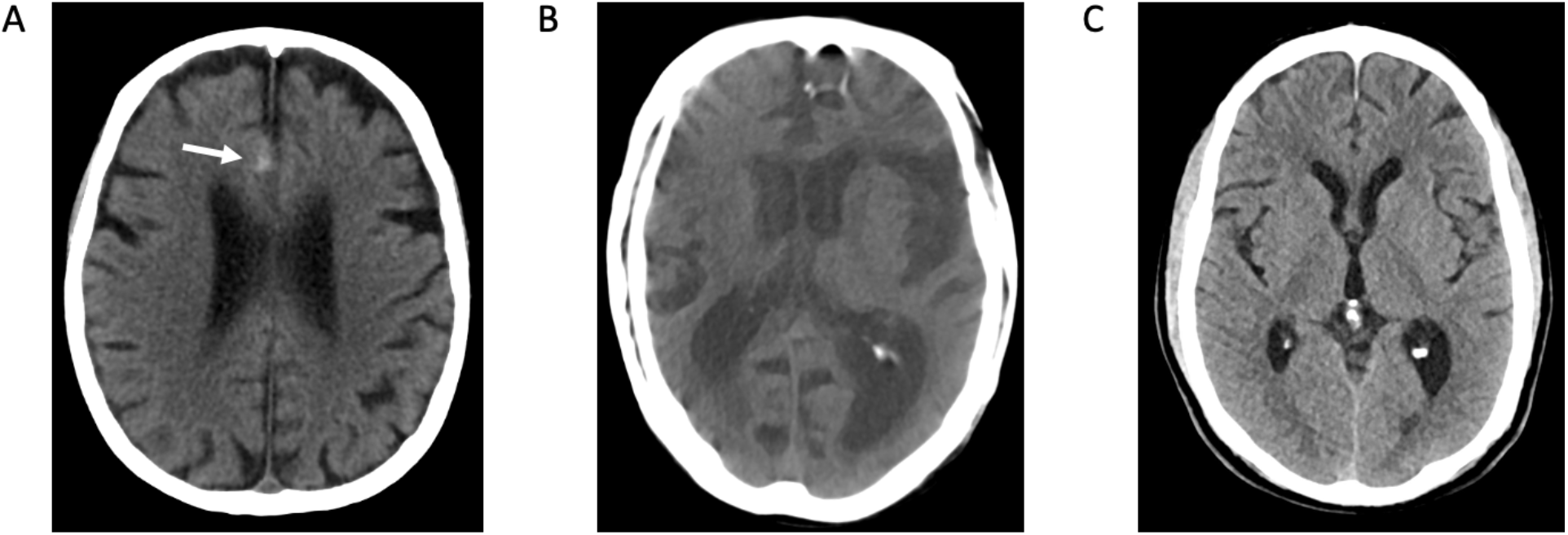
No Prediction cases. **A:** Trace acute subarachnoid hemorrhage along the right anterior cingulate gyrus. **B:** Noncontrast head CT degraded by motion artifact. **C:** Normal noncontrast head CT.

Interestingly, the NP group included the highest number of scans with artifacts, 19, and postoperative changes, 33. Forty-nine of the scans were normal. The rates of admission, neurosurgical intervention, and 30-day mortality were 53.4% (44.9-61.7), 12.0% (7.5-18.7), and 9.0% (5.2-15.2), respectively. The rates of admission and neurosurgical intervention were significantly higher for the IC+ cohort compared to NP with the odds ratios of 2.57 (1.32-4.97, P = 0.005) and 3.92 (1.88-8.19, P <0.001), respectively. When compared to the IC- cohort, the NP cohort had significantly higher rates of admission and neurosurgical intervention with the odds ratios of 1.50 (1.03-2.18, P = 0.033) and 3.03 (1.59-5.77, P < 0.001), respectively. The rate of 30-day mortality of the NP was not significantly different compared to the rate for the IC+ cohort (odds ratio 1.06 (0.38-2.97), P = 0.910) but was significantly higher than the rate for the IC- cohort (odds ratio 3.22 (1.54-6.76), P = 0.002).

There were 33 patients who had an acute, subacute, or age-indeterminate ischemic infarction. Twenty-eight patients were classified as IC- and 5 patients were classified as NP group.

## Discussion

Diagnostic ambiguity is common when a patient is evaluated for a new neurological symptom or after an event such as trauma. For example, symptoms such as a facial droop and speech difficulty may result from ischemia, hemorrhage, mass lesion and other pathologies. Even greater ambiguity occurs in patients who are unable to respond. Imaging is effective in narrowing the differential diagnosis, but imaging and image interpretation are also imperfect. Since time is of the essence, ML algorithms that quickly identify imaging abnormalities are promising new tools, but their inferences also have uncertainties that must be recognized and managed.

Following the suggestions by Kompa and colleagues(7), we incorporated uncertainty into an ML algorithm that analyzes head CT scans. The algorithmic system correctly identifies IC+ and excludes IC- scans with intracranial hemorrhage or other urgent intracranial abnormalities in most patients. The studies with high algorithmic uncertainty were placed in the NP group. This is a system that has utility as a prioritization tool in busy settings and may be especially useful to caregivers when radiologists are not available.

Transfer learning is routinely used in machine learning. In transfer learning, a network trained to solve one problem using one dataset serves as the starting point for training the network for a related problem using a completely different dataset. The first dataset provides domain adaptation and cuts down the training time for the second problem. By exploiting what has been learned in one setting helps to improve generalization in another setting. Our use of a network trained for ICH to detect other abnormalities represents a form of transfer learning that works by learning features that are common to both ICH and other abnormalities. Most likely our trained network learned certain features from the base dataset and inductively transferred them to other brain abnormalities. Our network performs well for ICH detection, but the process of training narrowed the model bias in a beneficial way where it became useful for other intracranial abnormalities such as tumors and other hemorrhage-like conditions. While our network’s performance on non-ICH conditions can be further improved by subjecting it to more specific training, its performance even without such training is remarkable.

### IC+ Scans

The IC+ cohort comprised about 10% of the test group and had a >90% positive predictive value for an intracranial hemorrhage or other urgent intracranial abnormality. Moreover, three-fourths of this group were admitted, over a third had a neurosurgical intervention and nearly a tenth died within 30 days. It is evident that an IC+ classification is a powerful biomarker, and those patients should be promptly assessed clinically, and their scans immediately reviewed by an imaging expert. In a setting where a radiologist is not available, an IC+ classification provides an important alert to caregivers and can prompt additional action to manage the patient. It is noteworthy that the even the false negative IC+ scans had imaging abnormalities, mostly encephalomalacia or artifacts.

### IC- Scans

The IC- cohort had a 94% negative predictive value for intracranial hemorrhage or other urgent intracranial abnormality. This classification identified patients with significantly lower rates of admission, neurosurgical intervention, and 30-day mortality. Nearly all the false-negative cases within this cohort were due to a small amount of intracranial blood. While triage to a lower priority for review of these scans may be justified, IC- scans still require review by trained interpreters to identify small bleeds.

### NP Scans

The patients whose scans were placed into the uncertain NP cohort had interesting characteristics. Of the 168 NP scans, 31% had image artifacts or post-operative changes and 29% were normal. However, nearly a third had intracranial hemorrhage or other urgent intracranial abnormalities. Thus, immediate review of these scans by a radiologist or trained interpreter of head CTs is warranted for these scans, making the reviewer aware of the 1 in 3 chances of the presence of an intracranial hemorrhage or other urgent intracranial pathology.

### Early Ischemic Stroke

The ML algorithm does not identify ischemic injury. Not a single case of ischemic infarction was classified as IC+. Acute, subacute, and age-indeterminate infarctions occurred in 27 patients with scans classified as IC-, and 5 patients assigned to the NP group. These results were expected because although ischemic stroke may produce severe neurological deficits, early ischemia may not create CT scan abnormalities that are detectable by even experienced neuroradiologists. It is thus not surprising that the ML algorithm presented here did not infer an abnormality in early ischemia patients. When imaging is needed to confirm ischemia, patients often undergo diffusion MRI. Our algorithm would not change this. However, progress has been made to train an algorithm to detect early cerebral ischemia. Our group has created such an algorithm and it is currently undergoing validation. We foresee aggregating these algorithms to provide a more thorough assessment of patients who present with symptoms that may suggest a stroke. Such a suite of algorithms would be especially valuable in settings where a neuroradiologist may not be immediately available.

### Potential clinical applications

The head CT ML algorithm described here performs sufficiently well to serve as a radiology assistant. It is a tool that can help radiologists manage busy imaging centers such as the emergency department. It might be extremely valuable when a radiologist is not available. Rapid evaluation of studies in urgent care environments is needed to properly manage patients, especially those with intracranial pathology. It is easily feasible for algorithm analyses to be available at the time that the scans appear on the image review station and be marked as IC+, IC- or NP. Since IC+ scans have a >90% probability of intracranial hemorrhage or other urgent intracranial pathology as well as one-third of the NP scans, the radiologist’s attention can immediately focus on the most impactful scans. The radiologist would also know that the IC- scans are usually normal, but that small hemorrhages may be present and may thus focus her diagnostic skills to identify subtle bleeds.

Other ML algorithms have been shown to reduce the turnaround time for the identification and interpretation of intracranial hemorrhage or other urgent intracranial abnormalities such as intracranial hemorrhage on head CTs.(19) However, many algorithms have the limitation of only prioritizing studies with a single intracranial abnormality such as hemorrhage, which can lead to inadvertent delays in the interpretation of studies with other intracranial abnormalities that require urgent treatment. For instance, in a clinical practice where an ICH detection algorithm is implemented for worklist prioritization, a study with a large non-hemorrhagic mass with herniation requiring immediate attention may be shifted to the lower end of the reading queue due to the absence of ICH. Our ML algorithm is more likely to expedite the identification of patients with more urgent needs for admission and intervention. This system also allows flexibility in additional types of clinical settings. For instance, practices that have a longer time interval between the study scan time to radiologist interpretation, such as outpatient imaging centers, may triage IC+ and NP cases to ensure more timely identification of an intracranial hemorrhage or other urgent intracranial abnormality, which could otherwise wait for some time on a radiology worklist before the images are seen by anyone. This algorithm would be especially valuable in emergency and urgent care centers where a radiologist may not be available when the patient is scanned.

### Shortcomings and Future Directions

The algorithm has shortcomings. Most importantly the algorithm was trained only on scans where hemorrhage was present or absent. Despite the absence of training using scans with other pathologies, the algorithm performance was surprisingly good but can be improved. The obvious path forward is to retrain the algorithm using scans with other pathologies, which we are doing. The decision rules used to define classification (i.e., IC+ are those with a predicted probability of at least 0.9 on 3 slices) were based on an empirical evaluation of a test data set but may not be optimal. Other thresholds may improve the performance of the algorithm and we will investigate this further after retraining the algorithm.

To reduce selection bias, 1000 consecutive cases that were processed by the algorithm were included in the study. While most of the studies were from the emergency department, a subset of cases included inpatient, intraoperative, and outpatient cases. However, these were also deemed urgent or emergent and were directed to the emergency neuroradiology service for interpretation. Finally, the algorithm was not used to assist in the real-time evaluation of imaging studies, so it remains uncertain how much of a clinical impact the algorithm would have in actual clinical practice.

## Conclusion

This ML algorithm that incorporates uncertainty can categorize head CT scans with or without intracranial hemorrhage or other urgent intracranial abnormalities with high predictive values and result in clinically relevant classification. It may help expedite patient triage and appropriate management in various clinical settings.

## Data Availability

All data produced in the present study are available upon reasonable request to the authors

